# 4D Ultrasound Localization Microscopy of Deep Cerebral Perforating Arteries for Intraoperative Neurosurgical Guidance

**DOI:** 10.64898/2026.02.06.26345454

**Authors:** Yichuang Han, Yasmin Sadigh, Luuk Verhoef, Luxi Wei, Sadaf Soloukey, Arber Demi, Paul Xing, Peter de Smalen, Arend Jan de Jong, Francesca De Carlo, Emma Gommers, Arnaud J.P.E. Vincent, Ruben Dammers, Johan G. Bosch, Pieter Kruizinga, Victor Volovici, Jason Voorneveld

**Affiliations:** BrainEcho Lab, Department of Neuroscience, Erasmus MC, Rotterdam, the Netherlands; Thorax Biomedical Engineering, Department of Cardiology, Erasmus MC, Rotterdam, the Netherlands; Department of Neurosurgery, Stroke Center, Erasmus MC, Rotterdam, the Netherlands; Center for Complex Microvascular Surgery, Erasmus MC, Rotterdam, the Netherlands; Department of Engineering Physics, Polytechnique Montréal, Montréal, Canada; Department of Anesthesiology, Erasmus MC, The Netherlands; Oldelft Ultrasound, Delft, the Netherlands; Department of Neurosurgery, Erasmus MC, Rotterdam, the Netherlands; Brain Tumor Center, Erasmus MC Cancer Institute, Erasmus MC, Rotterdam, the Netherlands; Signal Processing Systems, Delft University of Technology, Delft, The Netherlands

## Abstract

The human cerebral microvasculature is both essential for brain function and highly vulnerable, yet its *in-vivo* structure and local hemodynamics remain largely unexplored due to the lack of imaging techniques capable of resolving deep microvascular flow in humans. This limitation not only constrains fundamental neurovascular research, but poses life-altering risks during neurosurgery, where damage to small perforating arteries can have devastating neurological consequences. Specifically, the deep cerebral perforators, branching from the major trunks of the Circle of Willis, supply essential regions of the cerebral central core but remain beyond the resolution of current intraoperative imaging modalities. Here, we report a first in-human cohort study (10 patients) demonstrating the use of 4-dimensional ultrasound localization microscopy (4D-ULM) for the intraoperative visualization of cerebral microvascular anatomy and hemodynamics. In eight patients, 4D-ULM enabled volumetric mapping of deep perforators with sub-millimeter spatial resolution (∼140 µm) at depths reaching 7 cm. This approach revealed detailed flow patterns within the previously inaccessible deep vascular networks of the human brain. Our results open new opportunities for studying microvascular physiology and could enhance intraoperative decision-making by providing high-resolution hemodynamic data, paving the way for improved microsurgical precision in neurosurgical procedures.

## Introduction

One of the most challenging aspects of neurosurgery is the continuous need to accurately discern healthy blood vessels that vascularize eloquent areas of the brain from pathological vessels^1–8^. Of critical importance are the small (< 1 mm diameter), deeply situated perforating arteries that branch from the main vessels of the Circle of Willis to vascularize the central core of the brain (basal ganglia, thalamus, and internal capsule-Figure 1a)^9^. These areas are responsible for a myriad of cognitive, motor and sensory functions, contributing to all brain processes^9^.

**Figure 1:**
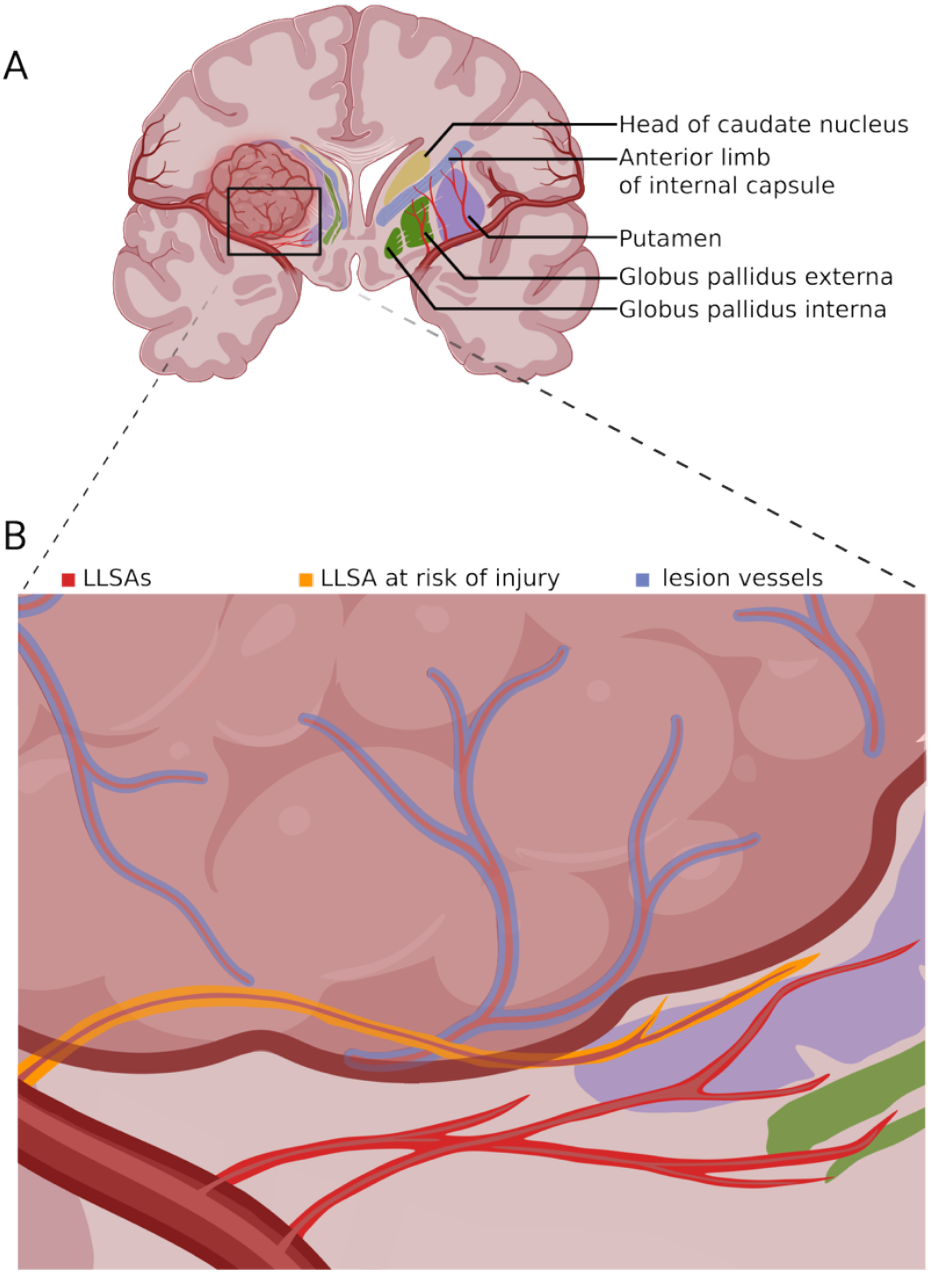
Schematic figure of deep cerebral perforating arteries and vascularized territory. A)Schematic illustration of the vascular territories supplied by deep cerebral perforating arteries. Lateral lenticulostriate branches vascularize the basal ganglia, thalamus, and internal capsule, regions critical for motor, sensory, and cognitive function. B) Representative pathologies encountered near these territories with perforator involvement. Intraoperative damage or occlusion of these arteries can cause severe neurological deficits; therefore, gross total extent of resection needs to be balanced with healthy vessel preservation to ensure maximal safe lesion resection.

In many lesions requiring neurosurgery, particularly brain neoplasms and vascular diseases, perforating arteries are often adjacent to or intertwined with pathological tissue, making them prone to unintended surgical injury (Figure 1b)^8,10,11^. Injury to perforating arteries may cause severe neurological deficits and significant impairments of postoperative functional outcome ^7,12^. Perforating arteries are “end-arteries”, without collateral circulation^9,13^, which implies that even the sacrifice of one submillimeter vessel may lead to permanent neurological deficits ^7,12^. Nevertheless, the surgeon strives to remove as much of the pathological tissue as possible, creating a delicate balancing act between preserving perforator integrity and maximal resection^1,2^. An intraoperative imaging technique capable of visualizing these submillimeter arteries remains a critical unmet need for improving surgical safety and postoperative outcomes. Given the size and anatomical complexity of perforating arteries, traditional imaging techniques lack the resolution to accurately visualize these vessels in real-time. As a result, surgeons face increased risk of inadvertently damaging or sacrificing critical arteries, leading to neurological deficits.

So far, only 7.0 Tesla Time-of-Flight Magnetic Resonance Angiography (TOF MRA) and photon-counting computed tomography angiography (PC-CTA) have demonstrated some ability in visualizing perforating arteries ^14–16^. Despite these advancements, high-resolution imaging still faces significant limitations in the context of intraoperative availability, and usage without disruption of the surgical workflow. Furthermore, these imaging modalities provide only static images, acquired preoperatively, which are prone to “brain-shift”: distortion of the vascular and parenchymal tissue during surgery^17^. This brain-shift can significantly alter the positioning of key vascular structures, making preoperative imaging data less reliable for real-time surgical guidance. While intraoperative use of MRA or CTA is logistically complex; the craniotomy creates a natural acoustic window that makes ultrasound imaging an ideal candidate for real-time surgical guidance.

A recent development in ultrasound imaging, “Ultrasound Localization Microscopy” (ULM), leverages ultrafast imaging ^18–22^ to localize and track intravenously injected microbubbles (< 10 µm diameter), achieving super-resolved visualization of microvascular structure and flow^23^. To date, most ULM work in the brain has been performed in two-dimensions (2D) and primarily in animal models^24–27^. Only a handful of studies have translated to humans^28–31^. Notably, recent transcranial experiments demonstrated that 2D ULM, acquired through the temporal bone window, can resolve perforating arteries supplying the basal ganglia^30^. In parallel, our group demonstrated the first intraoperative use of ULM and showed a proof-of-concept for three-dimensional (3D) ULM. This was achieved using ultrafast four-dimensional (“4D”) ultrasound imaging, which enables continuous, ultrafast volumetric acquisition over time (3D + time)^31^.

Extending ULM to 3-dimensions presented a formidable technical challenge. To coherently track microbubbles throughout the vascular tree, 4D imaging is required, at hundreds of volumes per second, which is not possible with conventional ultrasound hardware^32^. While several transducer designs have been explored for 3D-ULM in animal studies: including fully addressed ^33,34^, sparse ^35,36^, row-column^37,38^, and large-element probes^39^; they remain intraoperatively impractical due to high channel count requirements, sacrificed signal quality, or restricted fields of view. We achieve 4D ultrasound imaging using a miniaturized matrix-array probe (Adult 4D TEE, Oldelft Ultrasound) with embedded beamforming electronics (micro-beamforming) ^40^ connected with a research ultrasound machine (Vantage 256, Verasonics Inc). The probe’s compact footprint (∼1 cm^2^) allows placement within deep surgical corridors such as the Sylvian fissure or skull base -regions previously inaccessible to volumetric ultrasound. Combined with ultrafast contrast-enhanced acquisition and 3D super-localization algorithms, this system enables 3D-ULM, capturing the brains vascular networks with sub-millimeter resolution. Additionally, by temporally segmenting microbubble tracks into their cardiac phase^41^, we also demonstrate for the first time 4D-ULM, where the hemodynamics of individual blood vessels can be quantified and visualized at the same sub-millimeter scale.

In this study, we present the first intraoperative 4D-ULM of cerebral perforating arteries in ten neurosurgical patients. Using the miniaturized probe, we could image a 60° × 60° × 7 cm volume at 450 volumes per second, with a resolution of ∼137 µm. This allowed visualization of individual perforating arteries that were otherwise invisible on pre-operative PC-CTA. Beyond static morphology, the technique provided quantitative hemodynamic parameters, including blood-flow velocity, directionality, and pulsatility within individual vessels.

## Results

### Patient Characteristics

Ten patients diagnosed with brain lesions requiring surgical resection were enrolled (Table S1). The median age was 54 years (IQR 49-61), and seven patients were women (70%). In total, seven (70%) lesions consisted of skull base meningiomas, two (20%) were cavernomas, and one (10%) epidermoid cyst was included. Lesion locations were illustrated in Supplementary Appendix Figure S1. A pterional surgical approach was performed in six (60%) patients. Median lesion size was 25 mm (IQR 19-51). The targeted perforating arteries were the lateral lenticulostriate arteries (LLSA) in eight (80%) patients due to the lesion localization. In nine (90%) patients, preoperative or post-operative PC-CTA was acquired.

### Measurement Characteristics

In all cases, up to two intra-operative volumetric contrast-enhanced ultrasound acquisitions were obtained, maximally extending surgical time by 20 minutes (median duration of surgery 482 minutes, IQR 410-583). In total, sixteen acquisitions were obtained (Table S2). In twelve (75%) the probe was mounted on a modified surgical fixation arm (Trimano, Getinge) and tracked by the neuro-navigation system (Curve, BrainLab, Germany) ^42^ enabling precise co-registration of the ultrasound field with preoperative PC-CTA. For the other four (25%) acquisitions, the probe was inserted and held deeper manually after anterior clinoid process removal (Table S1), taking advantage of the probe’s small footprint.

After craniotomy, the ultrasound probe was positioned on the cortical surface and aligned to the target vascular territory using real-time volumetric power-Doppler imaging (PDI, Figure 2a).

**Figure 2:**
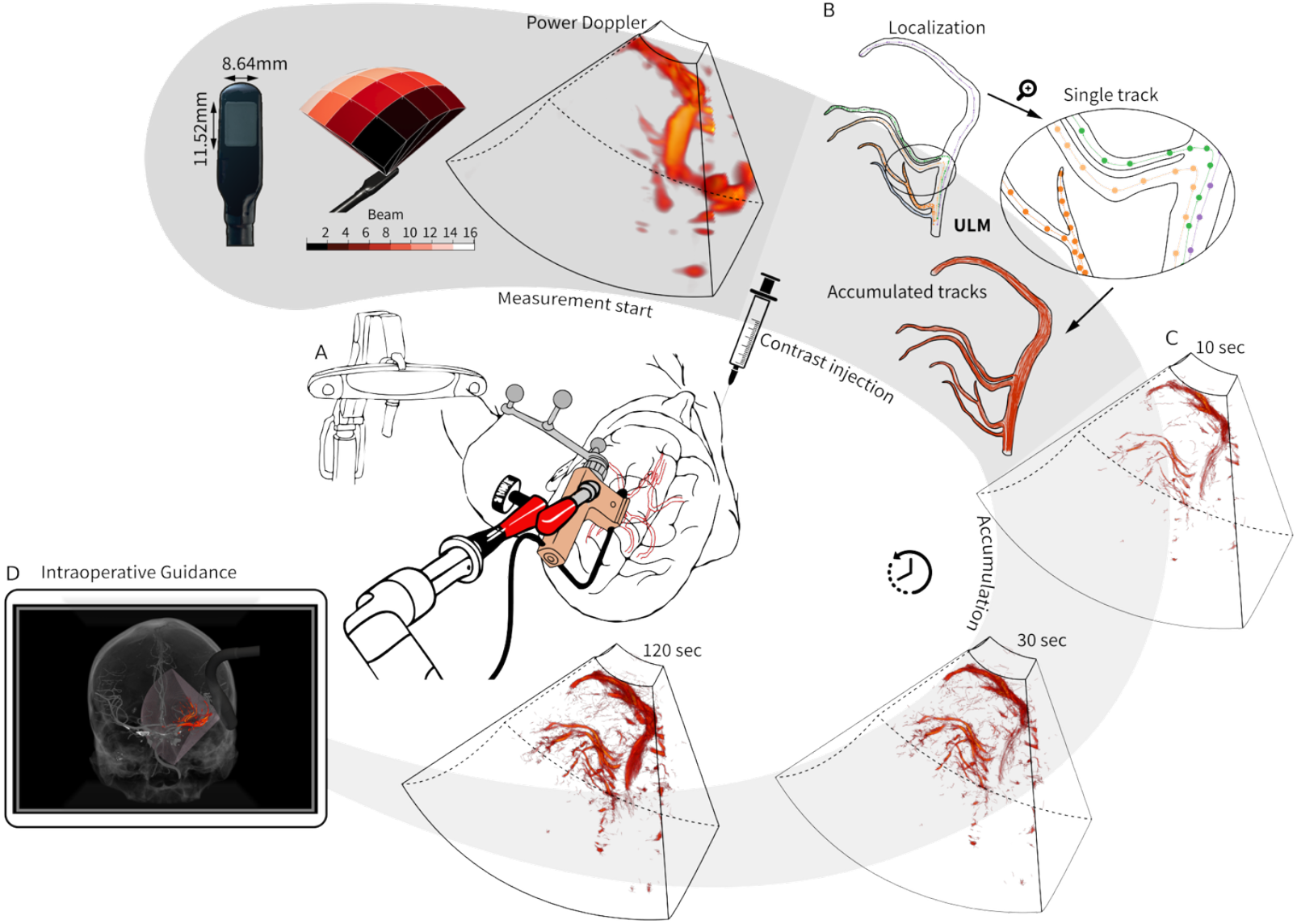
Intraoperative setup and ultrasound localization microscopy (ULM) pipeline. A) Schematic of the intraoperative imaging setup. The Oldelft Adult 4D TEE probe is mounted on a fixation arm and optically tracked by the BrainLab neuronavigation system for co-registration with preoperative photon-counting CTA. Real-time power Doppler imaging was used intraoperatively for acquisition guidance. B) ULM reconstruction pipeline. Beamformed contrast-enhanced data are SVD filtered before microbubble localization and tracking. The microbubble trajectories are then temporally accumulated to produce super-resolved vascular maps for three-dimensional rendering. C) Progressive vascular detail revealed after 10 s, 30 s, and 120 s of trajectory accumulation. D) ULM vascular map overlaid with pre-operative photon-counting CTA, illustrating intraoperative ULM with the patient’s anatomical context.

Each contrast-enhanced ultrasound acquisition started by intravenous injection of 0.4 mL bolus of SonoVue^®^ (Bracco, Italy), with simultaneous real-time PDI feedback while the raw channel data was streamed to disk for offline processing. Beamformed data underwent background removal to isolate microbubble signal^43,44^, after which each frame was motion-corrected for small tissue displacements, microbubbles were localized and tracked over consecutive frames (Figure 2b). Temporal accumulation of the microbubble tracks revealed progressive vascular detail over time: with major vessels emerging after 10 seconds of acquisition, while finer perforating arteries became discernible after 30 seconds, and stabilization in vascular detail beyond 120 seconds (Figure 2c).

Imaging perforating arteries with ULM was successful in eight (80%) subjects, and in 13 (82%) acquisitions obtained. In the two subjects where ULM was not feasible, acoustic coupling and imaging depth were limiting factors: in one case, sterile saline (required for acoustic coupling) drained rapidly from the cavity due to the angle of the surgical positioning, and in the other, tumor size (max. 60 mm) exceeded our preset imaging depth, putting the vessels of interest out of reach from our chosen imaging angle and maximum depth.

### ULM comparison with PC-CTA

To assess the spatial correspondence of intraoperative ULM, reconstructed vascular maps were co-registered with preoperatively (n=8) or postoperatively (n=1) obtained PC-CTA. Figure 3 presents eleven acquisitions from six patients for whom both BrainLab tracking registration and PC-CTA were available(patient 5 lacked PC-CTA; patient 1 lacked BrainLab tracking registration). Annotated vessel maps are provided in Supplementary Appendix Figure S2.

**Figure 3:**
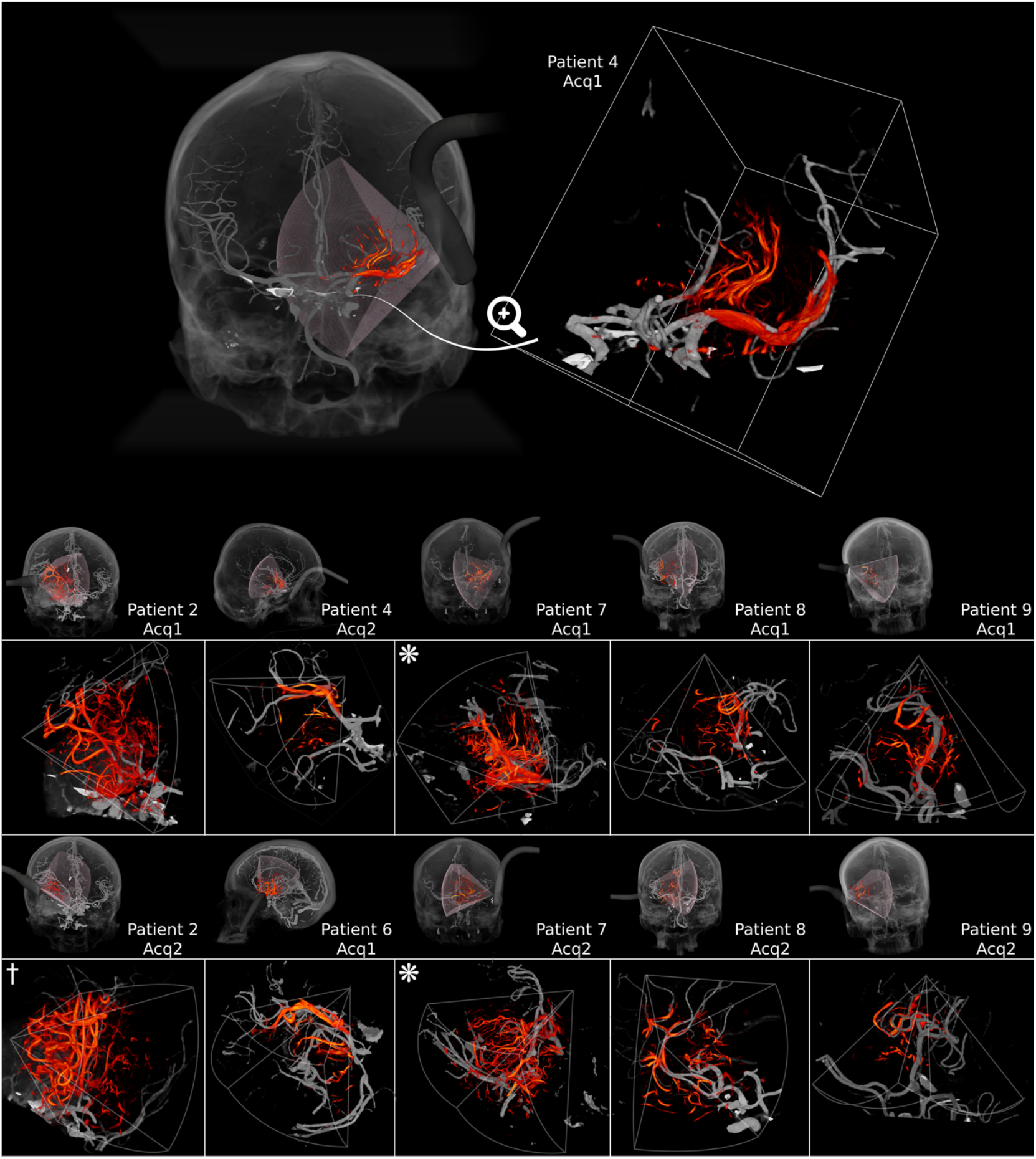
Morphological correspondence with PC-CTA and reproducibility of intraoperative 3D-ULM. Each panel shows a full-head PC-CTA rendering with the position of the ultrasound probe and its insonated field of view (transparent pink), co-registered to illustrate spatial correspondence between ULM volumes and patient anatomy. The adjacent magnified views display ULM-derived vasculature (red) overlaid on the corresponding PC-CTA (grey) within the insonated region. Patient and acquisition numbers are indicated on each panel. For P4-2 and P6-1, the probe was placed beneath the sphenoid wing. † Both pre-(P2-1) and post-resection (P2-2) ULM overlaid on preoperative PC-CTA. * Similarly, pre-resection (P7-1 & P7-2) ULM overlaid on postoperative PC-CTA. For P4-1, whole skull rendering was provided in Supplementary Appendix Video S1 and zoom-in vessel-tree rendering in Supplementary Appendix Video S2. Annotated vessel maps for all cases are provided in Appendix Figure S2.

Across all cases, overlay of ULM vascular maps with PC-CTA demonstrated strong spatial correspondence of the main trunks of the Circle of Willis (M1, M2, A1) and high-fidelity depiction of perforating arteries (Figure 3). ULM captured branching topology and was consistent with PC-CTA while revealing perforating arteries, which were not visible on PC-CTA. Of the sixteen intraoperative acquisitions, eleven yielded clear visualization of perforating arteries. In five acquisitions, perforating arteries were not identifiable due to no PC-CTA reference data (n = 1), absent spatial tracking (n = 1), poor acoustic coupling during intraoperative measurement (n =1), or insufficient depth of imaging due to a large tumor (n = 2). In all acquisitions requiring motion correction, slightly larger apparent vessel diameters were observed, most notably in patient 4 acquisition 2 (1.5 [IQR 1.4-1.5] mm vs. 0.8 [IQR 0.7-1.0]) mm across all acquisitions, likely reflecting motion-related blurring rather than true morphological differences (Table 1).

**Table 1:** ULM Morphological and Hemodynamic Measurements.

| PID | Acq | Morphological Measurements |  |  |  |  | Hemodynamic Measurements |  |
| --- | --- | --- | --- | --- | --- | --- | --- | --- |
|  |  | Stem | Branches | Length [mm] | Diameter [mm] | Tortuosity [-] | PI [-] | Velocity [mm.s<sup>-1</sup>] |
| 2 | 1 | 4 | 1 [1,2] | 27.4<br>(14.9, 41.8) | 0.7<br>(0.6, 0.7) | 1.3<br>(1.3, 1.4) | 0.41 (0.39,0.55) | 59.8<br>(49.9, 63.2) |
|  | 2 | 5 | 1 [1,2] | 28.6<br>(13.8, 37.3) | 0.9<br>(0.6, 0.9) | 1.3<br>(1.3, 1.5) | 0.59 (0.51,0.61) | 59.8<br>(53.5, 69.4) |
| 4 | 1 | 7 | 1 [1,3] | 16.4<br>(14.9, 27.2) | 0.7<br>(0.6, 0.8) | 1.3<br>(1.2, 1.4) | 0.86 (0.79,0.99) | 41.6<br>(37.5, 52.0) |
|  | 2* | 6 | 1 [1,2] | 10.6<br>(3.9, 12.6) | 1.5<br>(1.4, 1.5) | 1.2<br>(1.2, 1.3) | - | - |
| 6 | 1* | 6 | 2 [1,3] | 20.7<br>(10.6, 29.5) | 0.8<br>(0.6, 0.9) | 1.4<br>(1.3, 1.4) | 0.79 (0.43,0.81) | 61.0<br>(56.3, 71.0) |
| 7 | 1 | 5 | 4 [2,5] | 31.8<br>(29.1, 36.3) | 1.0<br>(0.7, 1.0) | 1.5<br>(1.4, 1.6) | 0.56 (0.51,0.60) | 49.9<br>(47.2, 55.5) |
|  | 2 | 7 | 4 [1,9] | 22.3<br>(16.2, 34.5) | 0.8<br>(0.7, 0.8) | 1.5<br>(1.3, 1.5) | 0.62 (0.59,0.66) | 55.2<br>(45.7, 58.4) |
| 8 | 1 | 5 | 2 [1,5] | 22.3<br>(13.8, 24.6) | 0.8<br>(0.8, 0.9) | 1.4<br>(1.3, 1.7) | 0.67 (0.54,0.72) | 34.3<br>(32.0, 41.2) |
|  | 2 | 7 | 2 [1,5] | 21.0<br>(19.0, 25.0) | 0.8<br>(0.6, 1.0) | 1.5<br>(1.3, 1.6) | 0.56 (0.30,0.65) | 35.6 (24.9, 41.9) |
| 9 | 1 | 6 | 1 [1,4] | 13.6<br>(3.4, 32.7) | 1.0<br>(0.8, 1.0) | 1.3<br>(1.2, 1.5) | 0.61 (0.60,0.65) | 42.7<br>(36.2, 48.9) |
|  | 2 | 2 | 2 [1,2] | 10.7<br>(3.2,18.2) | 0.4<br>(0.3, 0.4) | 1.4<br>(1.1, 1.6) | - | - |
| Total |  | 6 (5,7) | 2 (1,3) | 20.8<br>(11.6, 32.3) | 0.8<br>(0.7, 1.0) | 1.4<br>(1.3, 1.5) | 0.61 (0.50,0.75) | 48.9<br>(37.7, 57.3) |

1 Acq = Acquisition, PID = Patient identifier, PI = Pulsatility Index. Data displayed as Median [range] or (IQR).
\*Indicates acquisitions without probe fixation.

In certain cases, spatial overlap between pre- and post-operative datasets reflected expected surgical deformation. Specifically, patient 2 acquisition 2 represented a post-resection ULM volume aligned to pre-operative PC-CTA, while patient 7 acquisition 1 and patient 7 acquisition 2 represented pre-resection ULM aligned to post-operative PC-CTA. In these comparisons, spatial overlap decreased, likely caused by brain shift and local deformation following lesion debulking. These observations suggest that intraoperative ULM could serve as a real-time imaging tool to update preoperative imaging datasets to the intraoperative status, thereby maintaining anatomical accuracy despite brain shift during surgery.

### Perforator Morphology and Hemodynamics

Intraoperative volumetric ultrafast PDI was able to delineate the large vessels of the Circle of Willis (Figure 4a) such as Internal Carotid Artery (ICA), middle cerebral artery (MCA) M1 segment, anterior cerebral artery (ACA) A1 and A2 segments, in agreement with the estimated imaging resolution (2.1 × 2.7 × 1.3 mm) derived from the system’s point-spread function^40^. However, ULM post-processing of the same dataset unveiled an additional dense network of fine perforating arteries, specifically the LLSAs, branching from the M1 segment of the MCA (Figure 4b). A magnified view of a selected region (Figure 4c) illustrates the microvascular detail attainable with 3D-ULM, with a spatial resolution of 137 µm determined by Fourier shell correlation (Figure 4d), an order of magnitude finer than PDI.

**Figure 4:**
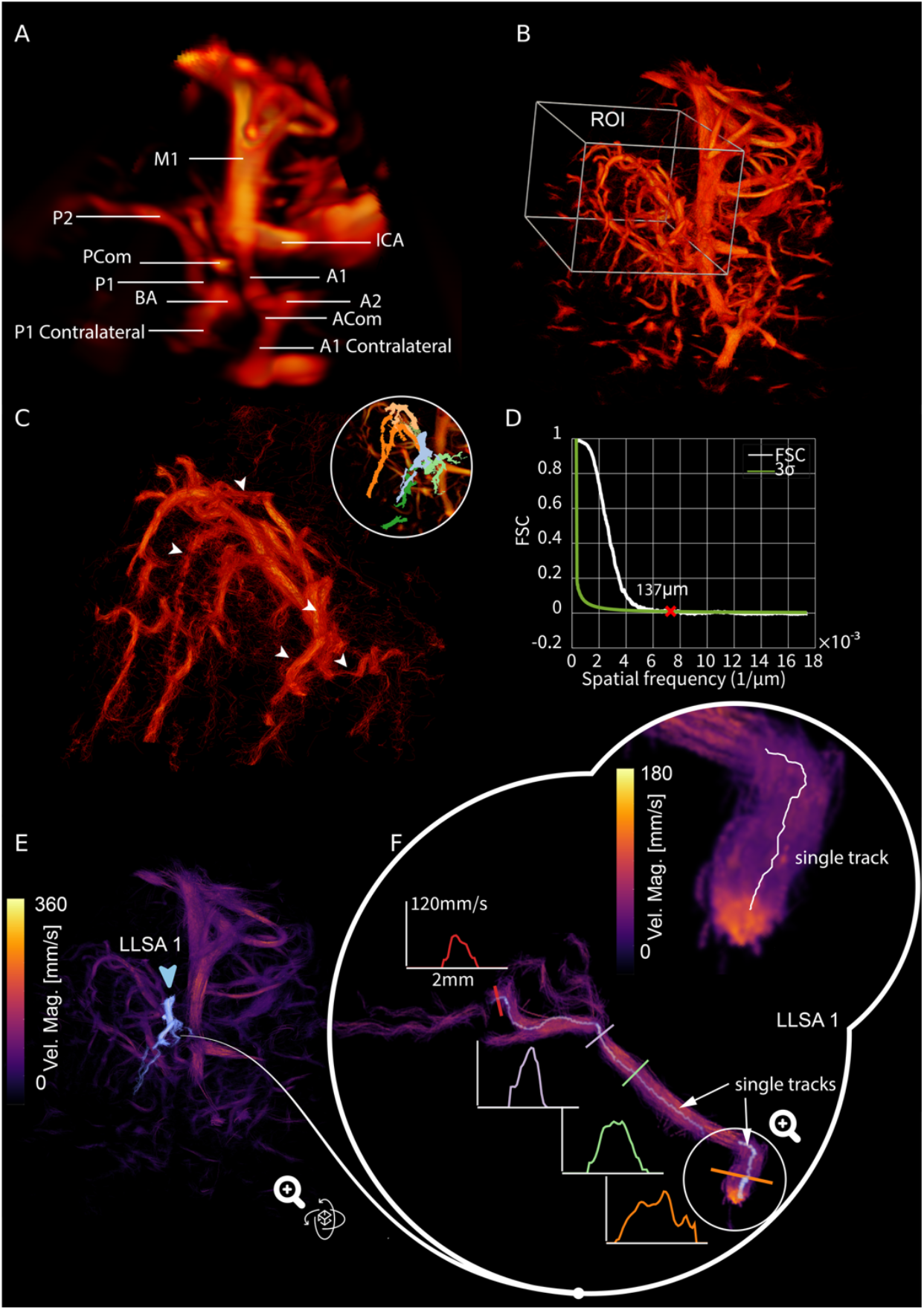
Structural and hemodynamic characterization of perforating arteries with intraoperative 3D-ULM. Unfolding levels of vascular detail: A) 3D Power Doppler image of the circle of Willis and its main branches. B) ULM, of the same region in A, reveals the lateral lenticulostriate arteries (LLSA) branching from the M1 trunk. **C)** Magnified region of interest from B showing sub-millimeter vessels (inset - individual perforator segmentations) resolved at 137 µm spatial resolution, as quantified by Fourier shell correlation shown in **D. E)** Average flow-velocity map of the same acquisition, showing flow deceleration from parent vessels to distal branches. **F)** Magnified flow-velocity view of one LLSA (marked in E), with cross-sectional velocity profiles showing progressive reductions in both vessel diameter and flow velocity from the stem to the distal end. Inset further zooms in on the vessel, showing the vessel cross section made from thousands of microbubble tracks (single track shown in white). M1, M2: segments of the middle cerebral artery. A1, A2: segments of anterior cerebral artery, P1, P2: segments of posterior cerebral artery. ICA: Internal carotid artery. BA: Basilar artery. ACom: Anterior communicating artery. PCom: Posterior communicating artery. Patient 20 - Acq 1 is shown.

Beyond morphology, 3D-ULM enables quantitative hemodynamic measurements of cerebral microcirculation. By aggregating individual microbubble tracks by their instantaneous velocity, a time-averaged flow-velocity map was obtained (Figure 4e). In a single LLSA (Figure 4f), four cross-sectional velocity profiles revealed a gradual narrowing of vessel diameter accompanied by a corresponding decline in flow velocity along the distal trajectory. The cross-sectional velocity distribution approximated a parabolic shape, consistent with laminar flow behavior in small arteries. Quantitative morphological parameters, including vessel diameter, length, and tortuosity, were comparable between subjects across different probe orientations (Table 1). A median of six perforator stems (IQR 5-7) were encountered, with individual perforating arteries having a median of two branches (IQR 1-3). A median length of 20.8 mm (IQR 11.6-32.3) of each individual perforating artery was recorded. Median vessel tortuosity was 1.4 (IQR 1.3-1.5). Previous 7T TOF MRA studies reported a mean diameter of 0.8-1.7 mm, whereas we found a median of 0.8 mm (IQR 0.7-1.0) in diameter across all cases (Table 1) ^45^.

Additionally, cardiac phase-resolved velocity mapping of individual vessels was achieved by aggregating track velocities across binned phases of the cardiac cycle ^46^ enabling vessel-specific characterization of perforator hemodynamics (4D-ULM). Individual velocity-time curves revealed distinct waveform patterns across patients (Figure 5). In patient 4 acquisition 1, perforating arteries displayed a pronounced systolic peak followed by a smaller diastolic crest, indicating an unusually pulsatile flow pattern in the deep brain compared to other patients, that may reflect increased arterial stiffness (Figure 5b). In contrast, patient 7 acquisition 2 had smoother single-peak waveforms, characteristic of more compliant vessels (Figure 5e,f), which was more like the rest of the cohort (Figure 5c,d).

**Figure 5:**
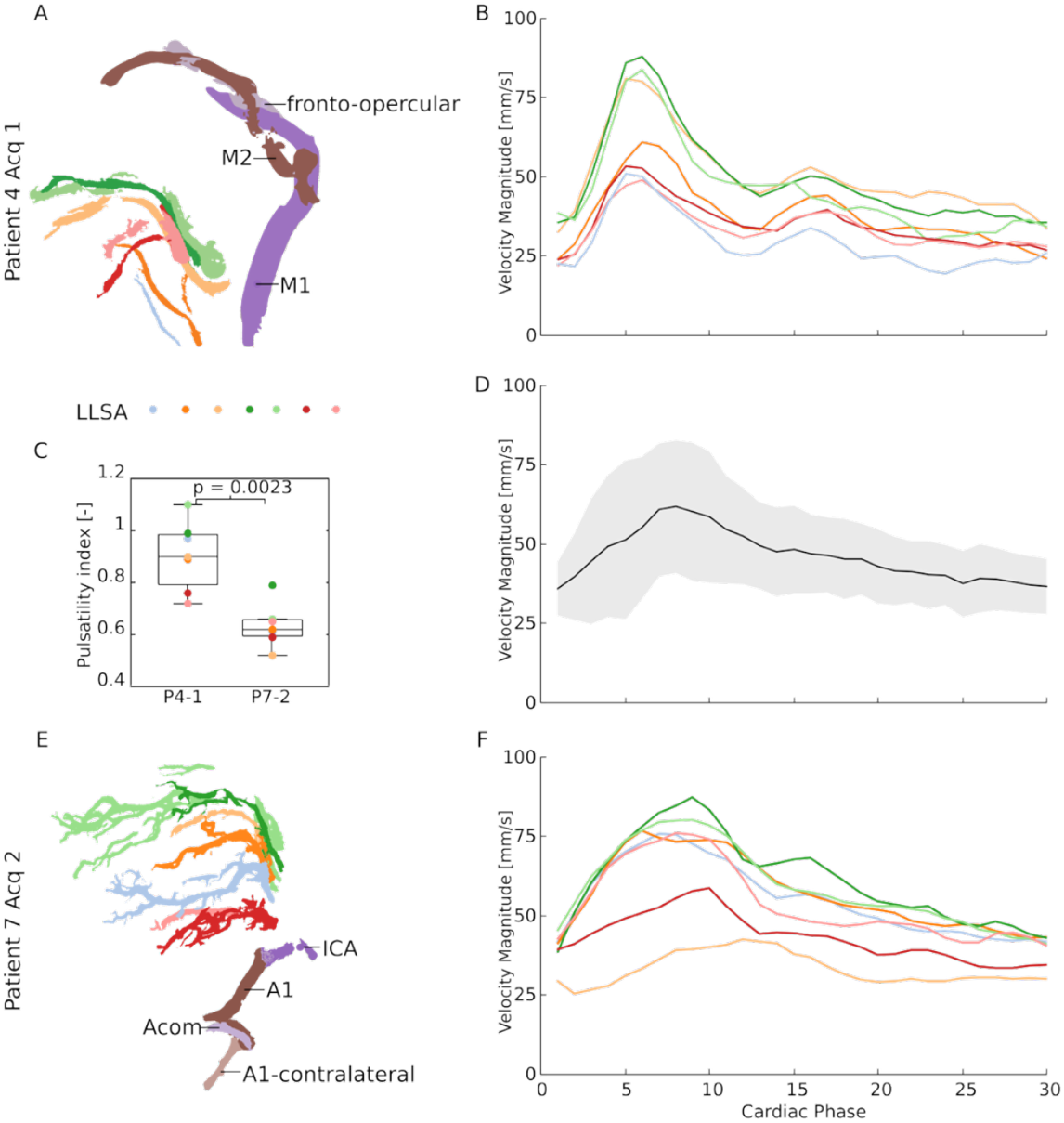
Quantification of vessel-specific hemodynamics in deep perforating arteries using 4D-ULM. The cardiac-phase resolved velocities of individual perforating arteries are shown for a patient (patient 4) with highly pulsatile flow during systole and diastole (**A&B**), in contrast to a more representative patient (patient 7) of the cohort (**E&F**), with only a systolic peak and gradual slowing over diastole. **C)** Wilcoxon rank-sum tests (un-paired, non-Gaussian) reveals significant differences (p<0.05) in pulsatility index of patients in **A** and **E. D)** Mean and standard deviation of cardiac-phase resolved velocities for all perforating arteries over the whole cohort. The overall mean peak systolic velocity was 69.1± 20.3 mm.s^-1^, with a mean PI of 0.64 ± 0.23.

Across all patients, the mean peak systolic velocity was 69.1± 20.3 mm.s^-1^, and the mean pulsatility index (PI) was 0.64 ± 0.23. The PI of perforating arteries measured in previous 7T TOF MRA studies consisted of a wide interval, ranging from a mean PI of 0.28 to 1.07 ^47–50^.The broad range in PI measurements could be explained by the anatomical and inter-group variations of perforating arteries.

These findings demonstrate that intraoperative ULM can quantify physiological variability close to findings from previous 7T TOF MRA measurements in microvascular dynamics, offering potential markers of local vascular dynamics, providing functional insight beyond structural visualization.

### Potential for Intraoperative Guidance

To illustrate the clinical applicability of ULM, pre- and post-resection imaging was performed in a patient with a large (maximum diameter 51 mm) sphenoid wing meningioma (patient 2, Figure 6a). The lesion displaced the MCA and LLSAs medially and posteriorly, compressing the insula, basal ganglia, and internal capsule.

**Figure 6:**
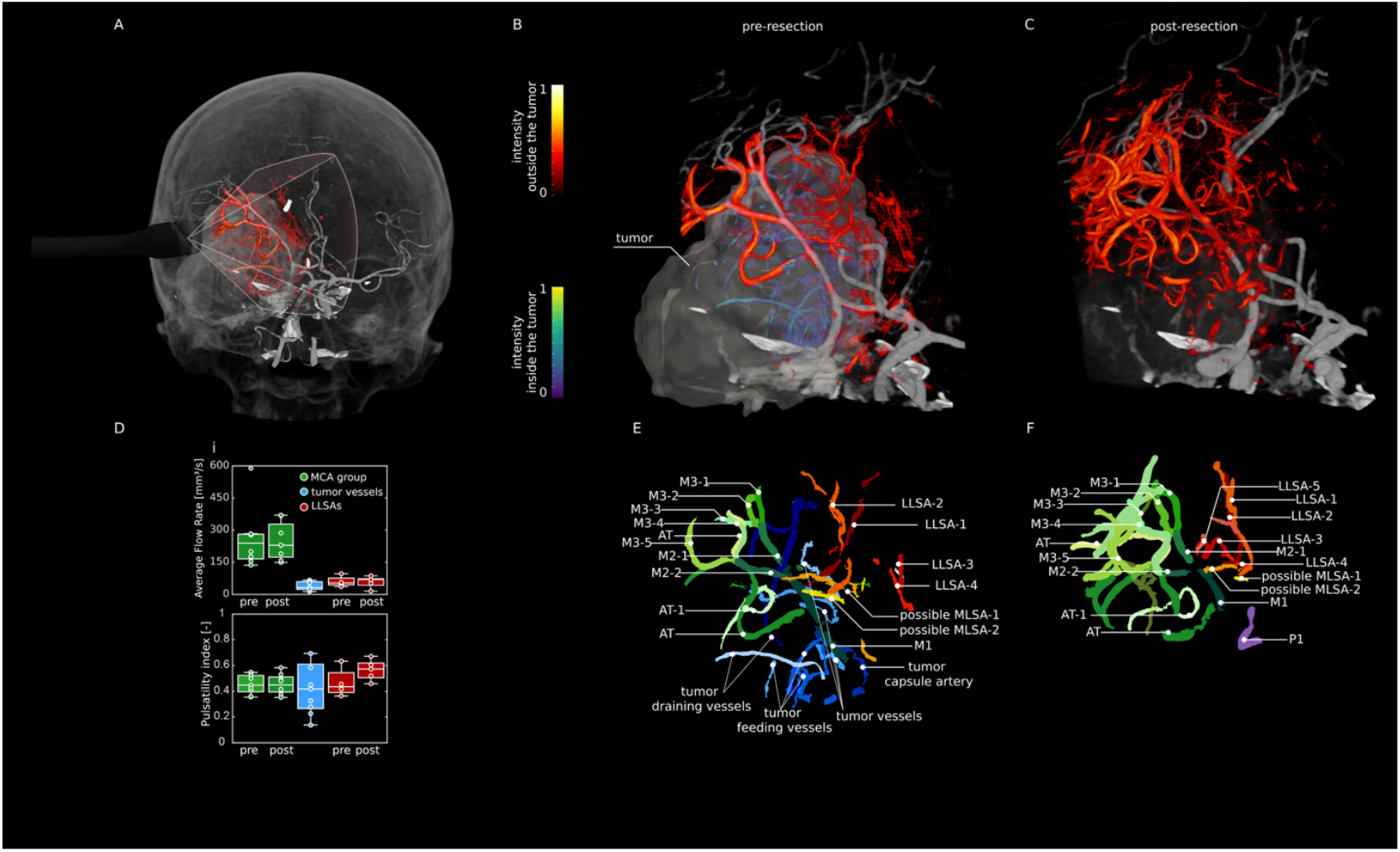
Intraoperative 3D-ULM of a large skull-base tumor in patient #2 before and after resection. The patient was diagnosed with a large (maximum diameter 51 mm) sphenoid wing meningioma. The lesion displaced the MCA and LLSAs medially and posteriorly, compressing the insula, basal ganglia, and internal capsule. A) Full-skull rendering of the pre-resection ULM acquisition co-registered PC-CTA, also available in Supplementary Appendix Video S3. B) Magnified view of A, with vessels inside and outside the lesion (segmented from preoperative MRI) rendered in distinct color maps. C) Post-resection ULM co-registered with pre-operative CTA. D) Box plots of flow rate and pulsatility index for each vascular group (MCA branches, LLSAs, and lesion vessels) annotated in E (pre-resection) and F (post-resection). E–F) Annotated vascular maps corresponding to pre- (B) and post-resection (C) acquisitions, rendered in the same orientation, also available in Supplementary Appendix Video S4 and S5. Large healthy vessels are shown in green tones, lesion-associated vessels in blue, and lenticulostriate arteries in red. M1, M2, M3: segments of middle cerebral artery. AT: anterior temporal artery. LLSA: lateral lenticulostriate arteries. MLSA: medial lenticulostriate arteries. P1: segment of posterior cerebral artery.

Using the pre-resection ULM, co-registered with preoperative PC-CTA, three vessel classes could be identified: 1) healthy arterial trunks (M2, M3, and anterior temporal segments); 2) LLSAs; and 3) a network of pathological lesion-associated feeding and venous vessels draining near the sphenoid wing of the skull base. Further co-registration with preoperative MRI enabled segmentation of the lesion volume and classification of all vessels in context of the tumoral border (Figure 6b,e).

Following gross-total resection, post-resection ULM confirmed preservation of parent MCA branches and possible LLSAs (Figure 6c,f), while lesion-associated arterial feeders and draining veins were no longer visible. Boxplots of flow rate and PI across vascular groups are shown in Figure 6d. Flow rates are highest in MCA branches (M2–M3 and anterior temporal segments) and substantially lower in LLSAs and lesion-associated vessels. The LLSAs exhibited slightly higher PI than the MCA branches, consistent with their smaller caliber and greater distal resistance. In contrast, the lesion-associated vessel group displayed markedly broader PI variability, as its composition is more heterogenous including pericapsular and feeding arteries and draining veins. These results highlight intraoperative ULM’s potential for providing surgeons with almost immediate feedback on both vascular preservation and resection completeness. With real-time post-processing and integration into surgical navigation systems, ULM could enable confirmation of intact brain perfusion and lesion devascularization, potentially reducing the risk of postoperative ischemia in complex skull-base lesion resections.

## Discussion

This study is the first demonstration of intra-operative 3D-ULM as a morphological and hemodyamic imaging tool, able to visualize brain microvasculature down to the diameter of cerebral perforating arteries up to 7 cm depth of the brain. By combining real-time 4D ultrasound imaging with microbubble localization and tracking, we achieved sub-millimeter (∼137 µm) vascular resolution. Integration with existing neuronavigation systems allowed for incorporation of the whole-brain anatomical context of MRI and PC-CTA. ULM was processed offline in this study, largely due to computational requirements. However, the localization and tracking algorithms are highly parallelizable, making them ideal candidates for GPU acceleration in the future. The physical constraint for timely feedback from ULM is how long it takes to accumulate sufficient microbubble tracks, which we demonstrated to be around 30 seconds to image the perforating arteries. In a case harboring a large sphenoid wing lesion, we compared the pre- and post-resection hemodynamic measurements of pathological versus healthy vessels. We found the lesion-associated vessels to be absent post-resection, confirming the complete devascularization of the lesion. On the other hand, the velocity in the healthy MCA branches (M2-M3) remained the same as preoperative status, confirming the preservation of these vital structures during surgery. Additionally, we found that LLSAs had a higher PI than MCA branches and pre-resection and post-resection measurements showed a slight reduction in PI within the LLSAs, possibly reflecting improved blood flow after lesion removal. These findings further underscore the potential of intraoperative ULM to provide real-time feedback on both vascular preservation and resection completeness.

An important factor enabling intraoperative application of 3D-ULM is the compact matrix-array probe used in this study. Achieving 500 volumes per second with conventional probes is challenging: the number of transducer elements and corresponding radiofrequency channels increase quadratically from linear arrays (used in 2D imaging) to matrix arrays (required for 3D). Even a relatively small 64 × 48 element matrix would demand 3,072 parallel receive channels, far beyond the limits of current ultrasound systems. This limitation has been overcome by using microbeamforming technology, which permitted full aperture control over 3072 elements, 3x more than any fully sampled matrix study with ULM^32^, while still being addressable by a single ultrasound system, requiring only 192 receive channels.

Secondly, its small footprint allowed access through narrow craniotomies and deep surgical corridors, including the skull base (Figure 3, patient 4 acquisition 2 and patient 6 acquisition 1), where larger volumetric probes or even linear arrays (2D imaging) cannot be positioned. Looking ahead, the development of smaller, or even wireless, probes that could remain in place during surgery (potentially incorporating electromagnetic or ultrasound-based tracking systems independent of line-of-sight constraints) could enable continuous microvascular monitoring without impeding surgical access. Such evolution would transform intraoperative ULM from an intermittent imaging technique into a persistent physiological monitor, providing real-time feedback on vessel patency and brain perfusion throughout the operation.

Thirdly, ULM showed the ability to provide both morphological (3D-ULM) and hemodynamic (4D-ULM) information at the level of individual vessel segments (Figure 5). Furthermore, by binning velocities into their respective cardiac phase, vessel-specific flow waveforms could be reconstructed (Figure 6), revealing variability in vascular compliance and microvascular resistance across vessels. Although these observations are preliminary and limited by sample size, they highlight ULM’s potential to quantify physiological variability in cerebral microcirculation.

Beyond technical feasibility, this work establishes ULM as a new paradigm for intraoperative vascular imaging. For decades, knowledge of deep perforating artery morphology and physiology has depended on *ex vivo* studies or indirect angiographic inference. The present results provide direct, quantitative access to these vessels *in vivo* and *in situ*, under surgical conditions, bridging the long-standing gap between microanatomy and intraoperative visualization. The ability of ULM to measure cerebrovascular flow is a critical advancement for neurosurgical practice, particularly in the context of complex cases with deep vascular involvement. Currently, intraoperative tools to assess blood flow in deep cerebral vessels remain severely limited ^51,52^ which poses a challenge for defining normal flow across different vascular territories and identifying areas of altered blood flow dynamics intraoperatively. Our findings suggest that the ULM derived PI and velocity measurements can provide insights into cerebrovascular hemodynamics.

While this study utilized offline processing of ULM data, the potential for continuous real-time monitoring in the future could significantly enhance intraoperative decision-making. Real-time monitoring of PI would enable neurosurgeons to more accurately identify areas of hypoperfusion and evaluate the adequacy of revascularization efforts beyond solely blood flow measurement ^53–55^. For example, in cases where hypoperfusion or compromised flow is detected intraoperatively, surgeons could use this data to assess the need for additional interventions like flow augmentation procedures ^20^. This type of real-time feedback could lead to more personalized surgical strategies and timely intraoperative adjustments. Furthermore, the integration of continuous monitoring could help identify subtle changes in vascular integrity that might not be apparent through traditional imaging, providing confirmation of preserving critical vascular structures such as perforating arteries and main arterial trunks.

Current intraoperative imaging modalities, such as Indocyanine Green video-angiography (ICG VA) and intraoperative digital subtraction angiography (DSA), have significant limitations that can affect their use in complex neurosurgical procedures ^56,57^. ICG VA provides real-time, visual confirmation of blood flow using fluorescence ^58–60^. It is especially useful for evaluating superficial vessels (laying in the scope of the microscope’s field) and ensuring patency of large vessels, while being unable to visualize the deeper regions of the vasculature ^61,62^. Intraoperative DSA, regarded as the standard imaging modality for evaluation of cerebrovascular anatomy, offers high-resolution imaging of both large and small vessels while detecting pathological changes in blood flow ^63^. However, obtaining DSA images is an invasive procedure, requiring catheter insertion and contrast injection. Furthermore, intraoperative DSA is time-consuming and disrupts the surgical flow and is unfit for continuous monitoring during surgery due to contrast-agent usage.

In contrast, ULM with its potential for continuous real-time monitoring, could overcome many of these limitations by providing both high spatial and temporal resolution of vascular structures, including deep, small, and complex vascular territories. Continuous ULM monitoring in future neurosurgical procedures could offer a highly precise, objective approach to managing cerebrovascular flow dynamically, improving intraoperative guidance, and potentially optimizing patient outcomes in high-risk surgeries involving lesions reaching complex vascular territories. To achieve this, future development should focus on enhancing the resolution and speed of microbubble tracking, enabling high-fidelity imaging of deep vascular structures in complex anatomical regions, and improving the portability and accessibility of ULM systems.

Despite the promising performance of ULM reflected by our results, several limitations should be considered. First, the flexibility of the miniaturized probe raised a limitation of our optical neuronavigation tracking system, which was unable to track when the probe is placed beyond the line of sight of the infrared cameras. Achieving proper probe registration ensures that the ULM data can be correlated to pre-operative imaging. In this study, six out of eight patients had successful registration, which allowed for the integration of ULM-derived vasculature with the anatomical context. In particular, patients 1 and 5 faced challenges that impacted registration (no Brainlab tracking in case 1 and no PC-CTA in case 5), which underscores the importance of having both adequate tracking and comprehensive pre-operative imaging for successful registration.

Second, a limitation of the bolus microbubble injection method used, is the large variation in microbubble density over time. Using a continuous infusion pump and ECG-synchronized acquisitions could enable monitoring of hemodynamics over longer periods, revealing cycle-to-cycle variations in flow pulsatility and perfusion that may reflect dynamic changes in vascular compliance or autoregulation. Future studies should investigate the integration of continuous infusion techniques with real-time monitoring, potentially enhancing the precision of hemodynamic measurements and enabling a more accurate assessment of vascular dynamics throughout the whole procedure.

Another limitation of the current study is the length of the vessels imaged. In many acquisitions, we found that certain segments, particularly the origin of the LLSA (start of branch from parent vessel), were missing. This contrasts with images from 7T TOF MRI, which typically provides more comprehensive coverage of vascular segments. However, the diameter and hemodynamic characteristics of the vessels that we were able to image with ULM are comparable to those observed with 7T MRI in the literature. Future research should refine ULM algorithms to enhance robustness to the dynamic range of microbubble densities and velocities present in the region, with optimization in both microbubble localization and tracking.

Intraoperative 3D-ULM enables direct visualization and quantification of deep cerebral perforating arteries in humans, combining vessel morphology and hemodynamics at a resolution previously reserved for microscopy only. These results open the door to new avenues for studying human cerebrovascular anatomy *in-vivo* and set the stage for translation of ULM toward continuous real-time neurosurgical guidance.

## Methods

### Patient Inclusion

Between February and May 2025, consecutive patients who underwent craniotomies for complex intra-axial or extra-axial lesions were prospectively included for the intra-operative use of ULM for imaging the perforating arteries. Inclusion criteria were adult (> 18 years) patients harboring brain lesions near the central core, defined as the basal ganglia, internal capsule, the thalamus, and brain stem. Patients with contra-indications to the administration of the contrast-agent Sonovue (Bracco Imaging, Italy) were excluded. The study protocol was approved by the institutional review board (Erasmus MC Medical Ethics Review Committee, MEC-2018-037) and was conducted in accordance with the Declaration of Helsinki. Written informed consent was obtained from all patients or their legal representatives at least 24 hours preoperatively. Upon patient inclusion, the strategies for intraoperative ULM imaging were discussed for eligible cases during a preoperative consensus meeting (YH, YS, JV, VV). The number and angles of acquisitions were discussed and predetermined based on the surgical approach and anatomical lesion characteristics, observed on preoperative MRI and PC-CTA.

### Perforating Artery Group Definitions

Perforating arteries arise from the main trunks of the Circle of Willis, which were defined as ‘parent vessels’. Lateral lenticulostriate arteries (LLSA) were defined as perforating artery groups arising from the first segment (M1) of middle cerebral artery (MCA)^9^. Medial lenticulostriate arteries (MLSA) were defined as perforating arteries arising from the first segment (A1) of anterior cerebral artery (ACA). Recurrent arteries of Heubner were defined as perforating arteries arising from the first of second segment (A1/A2) of ACA, with regard to anatomical variation. Insular arteries were defined as perforating arteries arising from the second (M2) segments of the MCA. For internal carotid artery (ICA), posterior cerebral artery (PCA), and anterior choroidal artery (AchA), perforating arteries were grouped and named after parent vessels.

### 4D Ultrafast Ultrasound

4D ultrasound imaging was performed with an Adult 4D TEE probe (Oldelft Ultrasound, the Netherlands), connected to a research ultrasound system (Vantage 256, Verasonics, USA). The probe has 3072 elements with a square pitch of 180 µm, resulting in an aperture of 11.5 × 8.6 mm. The 3072 elements are reduced to 192 receive channels using 4×4 element sub-aperture processing groups (“micro-beamforming”) on an application specific integrated circuit embedded (ASIC) behind the transducer matrix. The ASIC enables individual control of the transmit delays for all 3,072 probe elements. The acquisition sequence used has been described previously ^31,40^, but in brief, a wide-beam transmission sequence was used, where 4×4 beams, each with a ∼15°x15° opening angle were scanned over azimuth and elevation to cover a 60°x60° field of view. The transmit pulse had a center frequency of 3.6 MHz. For real-time acquisition guidance, volumetric Power Doppler Imaging (PDI) was produced using ensembles of 128 frames (Doppler frame rate ≈ 3.5 Hz), with GPU-accelerated delay-and-sum beamforming and singular value decomposition (SVD) thresholding-based clutter filtering. During acquisition, the raw IQ-modulated (50% bandwidth) channel data was streamed to disk for offline ULM processing.

### Intraoperative Contrast Enhanced Ultrasound Imaging Protocol

Patients were anesthetized, positioned, and craniotomies were performed according to local institutional guidelines. Ultrasound imaging commenced after bone flap removal. The ultrasound probe was enclosed in a sterile cover filled with ultrasound gel. The probe was positioned according the preoperatively determined strategy (extradural/intradural and superficial/deep placement) to capture the predefined region of interest (ROI) using a modified intraoperative surgical arm (Trimano, Getinge, Germany) with a probe holder (Figure 2) and tracked by a neuronavigation system (Curve, BrainLab, Germany). In cases where the probe contact site was situated deeply and could not be reached by the static probe holder (due to the size of the craniotomy), then the probe was held manually and therefore not tracked. Adequate acoustic coupling was maintained by continuous irrigation with 0.9% NaCl solution at the probe contact site, if necessary. Real-time volumetric PDI was used to position the probe and viewing angle. In all cases, a first non-contrast acquisition was performed (data not analyzed in this study)

At this point an intravenous bolus of 0.4 mL SonoVue was administered by the anesthesia care team, for microbubble tracking, followed by 10 mL 0.9% NaCl solution. Simultaneously, real-time PDI feedback was provided, while the raw channel data was streamed to disk for offline processing. Each acquisition lasted 210 seconds following the arrival of the contrast agent. The number of acquisitions using contrast varied between one to a maximum of four contrast boluses and was predefined as a part of the surgical strategy. The entire measurement extended the conventional surgical procedure by a maximum of 20 minutes.

### ULM Image Postprocessing Pipeline

Channel data were first reconstructed using GPU based delay-and-sum beamforming. The grid has a resolution of 0.94° in azimuth and elevation directions and 0.5 mm along axial axis. SVD filtering^43,44^ was then applied to suppress tissue clutter and isolate the microbubble signal, using an ensemble size of 384 frames. ULM was subsequently performed in a chunk-based manner, with each chunk corresponding to 384 frames using PALA toolbox^64^ extended to volumetric (3D) data. A few modifications were added, including lag-1 autocorrelation for noise suppression, 3D Gaussian fitting and noise thresholding for localization, a constant-acceleration state mode Kalman filter for bubble tracking^65^ and makima interpolation for continuous tracks ^66^.

For acquisitions in which the probe was manually held by the surgeon, motion correction was applied to compensate for hand-induced and physiological drift. Intra-ensemble motion was estimated using PDIs which were generated using an ensemble size of 16 frames. Using the first PDI of the ensemble as a reference, we calculated 3D rigid-body transformations (six degrees of freedom) to align subsequent volumes. Optimal parameters were determined by minimizing the voxel-wise mean squared error via the Levenberg-Marquardt algorithm (lsqnonlin function in MATLAB). The resulting transformation matrices were applied to the localized microbubble positions. For inter-ensemble motion correction, preliminary ULM maps were reconstructed for each ensemble, and rigid affine transformations were computed between consecutive reconstructions to estimate cumulative motion. A second-pass global registration was then performed, using the temporal midpoint of the sequence as a reference, to correct residual misalignments. The combined motion parameters were finally applied to the full set of microbubble trajectories to generate the motion-compensated 3D-ULM volume.

### Tracking with Kalman Filter

The constant-acceleration Kalman filter model ^65^ is defined as follows:

For every active track 𝒯 _*j*_ (*k*) we maintain a 9-dimensional state vector

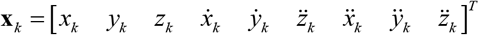

And extrapolate it to the next frame using a linear transition model (assuming a constant acceleration):

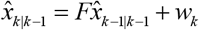

Where *w*_*k*_ is the process noise, modeled as Gaussian noise with covariance *Q*. And the state transition *F* is defined as:

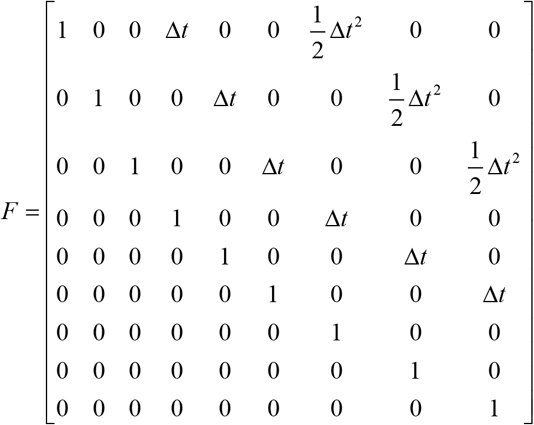

Where Δ_*t*_ is the time step between frames. The predicted position *z*_*k*_ is:

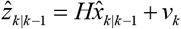

Where *H* is the observation matrix and ^*v*^_*k*_ is the measurement noise, modeled as Gaussian noise with covariance *R* .

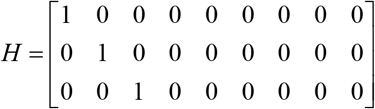

The Kalman state covariance will be updated correspondingly:

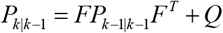

The Kalman gain is computed as:

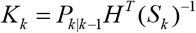

Where *S*_*k*_ = *HP*_*K* | *K-1*_ *H*^*T*^ + *R* is the innovation covariance.

The Mahalanobis-distance was used to construct the cost function for global nearest neighbor assignment:

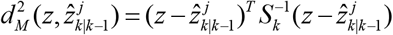

### Resolution estimation using Fourier shell correlation

The effective spatial resolution of the ULM reconstructions was quantified using Fourier shell correlation (FSC)^67^. The tracked microbubble trajectories were divided into two statistically independent subsets by temporal interleaving of chunks (odd versus even chunks). The two ULM density volumes were reconstructed from each subset using identical processing parameters. The three-dimensional Fourier transforms of the two volumes were computed, and the FSC was calculated as the normalized cross-correlation of corresponding Fourier coefficients within concentric spherical shells in spatial frequency space. The resolution was defined as the inverse spatial frequency at which the FSC dropped below the 3σ information threshold^67^, indicating the spatial frequency beyond which the reconstructions no longer contained reliable shared information (Figure 4d).

### Vessel Segmentation and Anatomical Annotation

The vascular density maps were first smoothed with a Gaussian spatial filter (filter size 3 × 3 × 3, σ = 0.5, imgaussfilt3, Image Processing Toolbox, MATLAB 2024a) and then enhanced using a Hessian-based Frangi vesselness filter^61^ to emphasize tubular structures (Supplementary Appendix Figure S3). The resulting vesselness volume was binarized and skeletonized to obtain one-voxel-wide centerlines. Centerline segments, defined as connected paths between junctions or endpoints, were automatically extracted using a customized graph-based segmentation algorithm in MATLAB (MathWorks), and each segment was mapped back to the enhanced vessel map and color-coded by its label to generate an automatically segmented vascular map.

The vascular map was imported into 3D Slicer (https://www.slicer.org)^68^ for manual refinement performed by one author (YS), using a customized annotation toolbox. The toolbox provides a simple way to interact with the segments created which is accomplished through single key-presses to add, hide or group segments together, and finally export the final annotation results to a readable format to be re-processed in MATLAB. This process included identification of perforating artery parent vessel (if captured by ULM) as a reference for anatomical orientation, initial noise removal, identification of perforating artery segments, assigning vessel labels to disconnected perforating artery segments, matching disconnected perforating artery segments to form the entire vessel, and removing remaining noise or structures beyond the ROI. Disagreements regarding the identification of perforating arteries and parent vessels were resolved through discussion with the senior clinical author (VV).

Following annotation, perforating artery segment reconnection was performed to ensure continuity of individual vessels for further analysis. First, annotated skeleton segments belonging to the same vessel label were re-linked based on both geometric and intensity criteria. Specifically, for each pair of segment endpoints, the surrounding vascular density map was searched to detect potential connecting paths. If any gaps existed between endpoints and contained voxels with intensity values above 30% of the mean intensity of the corresponding vessel segments predefined density threshold (0.2 × averaged local density) and spatially aligned with the endpoints, the gap was filled manually to bridge the segments.

From this bridged skeleton, vessel masks were reconstructed directly from the original vascular density map. For each annotated vessel, the skeleton served as a centerline from which the vessel cross-section was “regrown” by applying a local density threshold (30% of averaged intensity value for this vessel segment) in the surrounding voxels. This procedure restored the full 3D vessel volume corresponding to each annotated perforating artery, enabling quantitative morphological and hemodynamic analyses.

### Multimodal registration

For acquisitions in which optical tracking was feasible, the ultrasound transducer was tracked using an optical navigation system in conjunction with the standard patient reference marker. This setup enabled spatial co-registration of the intraoperative ULM data with preoperative imaging modalities, including CTA and MRI, following the workflow described previously ^42^. To further refine alignment, translational and rotational adjustments were performed manually in 3D Slicer to optimize the correspondence between major anatomical landmarks and improve global registration accuracy.

In cases where optical tracking was not possible due to restricted working space within the craniotomy (precluded attachment of the tracking marker to the probe), registration was performed manually offline in 3D Slicer. Alignment of the ULM volume to the preoperative photon-counting CTA (PC-CTA) was achieved by iterative adjustment of translation and rotation parameters, while visually matching key vascular landmarks (e.g., the M1 segment of the MCA, the circle of Willis, and distinctive bifurcations). Final registration accuracy was confirmed through consensus review by Y.S. and V.V.

### ULM Analysis of Perforating Arteries

ULM feasibility for depicting perforating arteries was achieved when the obtained vascular maps from intraoperative acquisitions were suitable for morphological and hemodynamic analyses.

### Morphological analysis (3D-ULM)

For each annotated perforating artery, the following morphological parameters were measured: (i) stem diameter in mm, defined as the diameter of the main trunk originating from the parent vessel; (ii) vessel length in mm, measured from the stem’s origin to the most distal endpoint along the longest branch; (iii) number of branches, determined from the segmented skeleton; and (iv) tortuosity, calculated as the ratio between the total centerline length and the straight-line (Euclidean) distance from the stem’s origin to its distal endpoint.

### Hemodynamic analysis (4D-ULM)

For hemodynamic analysis, the annotated vessel mask from each acquisition was overlaid onto the corresponding B-mode images, limiting the analysis to pixels within vessel regions. All masked B-mode frames were concatenated into a single spatiotemporal matrix, to which SVD was applied. The resulting temporal components were used to identify and segment individual cardiac cycles ^46^. Velocity waveforms for each perforating artery were then extracted across cycles, and cycle averaging was performed to obtain a representative velocity profile. Pulsatility index (PI) was then calculated as:

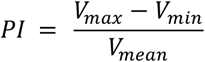

where *V*_*max*_, *V*_*min*_, and *V*_*mean*_ are the maximum, minimum, and mean velocities over the cardiac cycle, respectively.

### Statistical analysis

Descriptive statistics were calculated using MATLAB, with continuous variables summarized using medians and interquartile ranges (IQR) for non-normally distributed data and means with standard deviations (±) for normally distributed data.

## Data Availability

All data and relevant source code will be made available upon reasonable request from the corresponding author.

## Acknowledgements

The authors would like to thank the surgical and the anesthesia care team of Erasmus MC for facilitating the measurements.

## References

1. Gempt, J. et al. Postoperative ischemic changes following resection of newly diagnosed and recurrent gliomas and their clinical relevance. J. Neurosurg. 118, 801–808 (2013).

2. Gempt, J. et al. Postoperative ischemic changes following brain metastasis resection as measured by diffusion-weighted magnetic resonance imaging. J. Neurosurg. 119, 1395–1400 (2013).

3. Magill, S. T. et al. Postoperative diffusion-weighted imaging and neurological outcome after convexity meningioma resection. J. Neurosurg. 135, 1008–1015 (2021).

4. Strand, P. S., Sagberg, L. M., Gulati, S. & Solheim, O. Brain infarction following meningioma surgery— incidence, risk factors, and impact on function, seizure risk, and patient-reported quality of life. Neurosurg. Rev. 45, 3237–3244 (2022).

5. Strand, P. S. et al. Brain infarctions after glioma surgery: prevalence, radiological characteristics and risk factors. Acta Neurochir. (Wien). 163, 3097–3108 (2021).

6. De la Garza-Ramos, R. et al. Surgical complications following malignant brain tumor surgery: An analysis of 2002–2011 data. Clin. Neurol. Neurosurg. 140, 6–10 (2016).

7. van der Boog, A. T. J. et al. Occurrence, Risk Factors, and Consequences of Postoperative Ischemia After Glioma Resection: A Retrospective Study. Neurosurgery 92, 125–136 (2023).

8. Matsukawa, H. et al. Comprehensive analysis of perforator territory infarction on postoperative diffusion-weighted imaging in patients with surgically treated unruptured intracranial saccular aneurysms. J. Neurosurg. 132, 1088–1095 (2020).

9. Vogels, V., Dammers, R., van Bilsen, M. & Volovici, V. Deep Cerebral Perforators: Anatomical Distribution and Clinical Symptoms. Stroke 52, e660–e674 (2021).

10. Hou, Z. et al. Incidence of ischemic complications and technical nuances of arteries preservation for insular gliomas resection. Front. Surg. 9, (2022).

11. Isolan, G. R. et al. Avoiding vascular complications in insular glioma surgery – A microsurgical anatomy study and critical reflections regarding intraoperative findings. Front. Surg. 9, (2022).

12. Sadigh, Y. et al. Incidence and effect of supratentorial postoperative deep cerebral perforator territory ischemia in neurosurgical patients with intra-axial and extra-axial tumors. J. Neurosurg. 1–11 (2025) doi:10.3171/2025.7.JNS242861.

13. Ribas, E. C., Yağmurlu, K., de Oliveira, E., Ribas, G. C. & Rhoton, A. Microsurgical anatomy of the central core of the brain. Journal of Neurosurgery JNS 129, 752–769 (2018).

14. Harteveld, A. A. et al. High-Resolution Postcontrast Time-of-Flight MR Angiography of Intracranial Perforators at 7.0 Tesla. PLoS One 10, e0121051 (2015).

15. De Cocker, L. J. et al. Clinical vascular imaging in the brain at 7 T. Neuroimage 168, 452–458 (2018).

16. Okazaki, T., Niwa, T., Yoshida, R., Sorimachi, T. & Hashimoto, J. Visibility of Intracranial Perforating Arteries Using Ultra-High-Resolution Photon-Counting Detector Computed Tomography (CT) Angiography. Tomography 10, 1867–1880 (2024).

17. Gerard, I. J. et al. Brain shift in neuronavigation of brain tumors: A review. Med. Image Anal. 35, 403–420 (2017).

18. Tanter, M. & Fink, M. Ultrafast imaging in biomedical ultrasound. IEEE Trans. Ultrason. Ferroelectr. Freq. Control 61, 102–119 (2014).

19. Soloukey, S. et al. High-resolution micro-Doppler imaging during neurosurgical resection of an arteriovenous malformation: illustrative case. Journal of Neurosurgery: Case Lessons 4, (2022).

20. Niknejad, H. R., Huang, Y., Auricchio, A. M., van der Zwan, A. & Mischi, M. Intraoperative cerebral perfusion monitoring with ultrafast power doppler imaging. Sci. Rep. 15, 22003 (2025).

21. Imbault, M., Chauvet, D., Gennisson, J.-L., Capelle, L. & Tanter, M. Intraoperative Functional Ultrasound Imaging of Human Brain Activity. Sci. Rep. 7, 7304 (2017).

22. Soloukey, S. et al. Functional Ultrasound (fUS) During Awake Brain Surgery: The Clinical Potential of Intra-Operative Functional and Vascular Brain Mapping. Front. Neurosci. 13, (2020).

23. Errico, C. et al. Ultrafast ultrasound localization microscopy for deep super-resolution vascular imaging. Nature 527, 499–502 (2015).

24. Yi, H., Lowerison, M. R., Song, P. & Zhang, W. A Review of Clinical Applications for Super-resolution Ultrasound Localization Microscopy. Curr. Med. Sci. 42, 1–16 (2022).

25. Chen, Q., Song, H., Yu, J. & Kim, K. Current Development and Applications of Super-Resolution Ultrasound Imaging. Sensors 21, 2417 (2021).

26. Lowerison, M., Shin, Y. & Song, P. Super-Resolution Ultrasound Imaging: The Quest for Microvessels. Acoust. Today 20, 20 (2024).

27. Song, P., Rubin, J. M. & Lowerison, M. R. Super-resolution ultrasound microvascular imaging: Is it ready for clinical use? Z. Med. Phys. 33, 309–323 (2023).

28. Demené, C. et al. Transcranial ultrafast ultrasound localization microscopy of brain vasculature in patients. Nat. Biomed. Eng. 5, 219–228 (2021).

29. Schwarz, S. et al. Ultrasound Super-Resolution Imaging of Neonatal Cerebral Vascular Reorganization. Advanced Science 12, 1–13 (2025).

30. Denis, L. et al. Transcranial ultrasound localization microscopy in moyamoya patients using a clinical ultrasound system. Theranostics 15, 4074–4083 (2025).

31. Wei, L. et al. Intraoperative ultrasound localization microscopy of human brain tumors and arteriovenous malformations. Preprint at 10.1101/2025.09.19.25335978 (2025).

32. Denis, L., Chabouh, G., Heiles, B. & Couture, O. Volumetric Ultrasound Localization Microscopy. IEEE Trans. Ultrason. Ferroelectr. Freq. Control 71, 1643–1656 (2024).

33. Heiles, B. et al. Volumetric Ultrasound Localization Microscopy of the Whole Rat Brain Microvasculature. IEEE Open Journal of Ultrasonics, Ferroelectrics, and Frequency Control 2, 261–282 (2022).

34. Xing, P. et al. 3D ultrasound localization microscopy of the nonhuman primate brain. EBioMedicine 111, 105457 (2025).

35. Wei, L. et al. High-Frame-Rate Volumetric Porcine Renal Vasculature Imaging. Ultrasound Med. Biol. 49, 2476–2482 (2023).

36. Robin, J., Ozbek, A., Reiss, M., Dean-Ben, X. L. & Razansky, D. Dual-Mode Volumetric Optoacoustic and Contrast Enhanced Ultrasound Imaging With Spherical Matrix Arrays. IEEE Trans. Med. Imaging 41, 846–856 (2022).

37. Jensen, J. A. et al. Anatomic and Functional Imaging Using Row–Column Arrays. IEEE Trans. Ultrason. Ferroelectr. Freq. Control 69, 2722–2738 (2022).

38. Wu, A. et al. 3D transcranial Dynamic Ultrasound Localization Microscopy in the mouse brain using a Row-Column Array. IEEE Trans. Biomed. Eng. 1–12 (2025) doi:10.1109/TBME.2025.3598693.

39. Haidour, N. et al. Multi-lens ultrasound arrays enable large scale three-dimensional micro-vascularization characterization over whole organs. Nat. Commun. 16, 9317 (2025).

40. Verhoef, L. et al. Miniaturized Four-Dimensional Functional Ultrasound for Mapping Human Brain Activity. Preprint at 10.1101/2025.08.19.25332261 (2025).

41. Ghigo, N. et al. Dynamic Ultrasound Localization Microscopy Without ECG-Gating. Ultrasound Med. Biol. 50, 1436–1448 (2024).

42. Soloukey, S. et al. Human brain mapping using co-registered fUS, fMRI and ESM during awake brain surgeries: A proof-of-concept study. Neuroimage 283, 120435 (2023).

43. Baranger, J. et al. Adaptive spatiotemporal SVD clutter filtering for Ultrafast Doppler Imaging using similarity of spatial singular vectors. IEEE Trans. Med. Imaging 0062, (2018).

44. Demene, C. et al. Spatiotemporal Clutter Filtering of Ultrafast Ultrasound Data Highly Increases Doppler and fUltrasound Sensitivity. IEEE Trans. Med. Imaging 34, 2271–2285 (2015).

45. Schnerr, R. S. et al. Pulsatility of Lenticulostriate Arteries Assessed by 7 Tesla Flow MRI—Measurement, Reproducibility, and Applicability to Aging Effect. Front. Physiol. 8, (2017).

46. Ghigo, N. et al. Dynamic Ultrasound Localization Microscopy Without ECG-Gating. Ultrasound Med. Biol. 50, 1436–1448 (2024).

47. Arts, T. et al. Velocity and Pulsatility Measures in the Perforating Arteries of the Basal Ganglia at 3T MRI in Reference to 7T MRI. Front. Neurosci. 15, (2021).

48. Bouvy, W. H. et al. Assessment of blood flow velocity and pulsatility in cerebral perforating arteries with 7-T quantitative flow MRI. NMR Biomed. 29, 1295–1304 (2016).

49. Geurts, L. J., Zwanenburg, J. J. M., Klijn, C. J. M., Luijten, P. R. & Biessels, G. J. Higher Pulsatility in Cerebral Perforating Arteries in Patients With Small Vessel Disease Related Stroke, a 7T MRI Study. Stroke 50, 62–68 (2019).

50. Meijs, T. A. et al. Assessment of aortic and cerebral haemodynamics and vascular brain injury with 3 and 7 T magnetic resonance imaging in patients with aortic coarctation. European Heart Journal Open 3, (2023).

51. Tahhan, N. et al. Intraoperative cerebral blood flow monitoring in neurosurgery: A review of contemporary technologies and emerging perspectives. Neurochirurgie 68, 414–425 (2022).

52. Gulino, V. et al. The Use of Intraoperative Microvascular Doppler in Vascular Neurosurgery: Rationale and Results—A Systematic Review. Brain Sci. 14, 56 (2024).

53. Amin-Hanjani, S., Alaraj, A. & Charbel, F. T. Flow replacement bypass for aneurysms: decision-making using intraoperative blood flow measurements. Acta Neurochir. (Wien). 152, 1021–32; discussion 1032 (2010).

54. Amin-Hanjani, S., Meglio, G., Gatto, R., Bauer, A. & Charbel, F. T. The utility of intraoperative blood flow measurement during aneurysm surgery using an ultrasonic perivascular flow probe. Neurosurgery 58, ONS–305-12; discussion ONS-312 (2006).

55. Amin-Hanjani, S. et al. The cut flow index: an intraoperative predictor of the success of extracranial-intracranial bypass for occlusive cerebrovascular disease. Neurosurgery 56, 75–85; discussion 75-85 (2005).

56. Fritch, C., Church, E. & Wilkinson, D. A. Advances in Intraoperative Imaging for Vascular Neurosurgery. Neuroimaging Clin. N. Am. 34, 261–270 (2024).

57. Marbacher, S. et al. Comparison of 3D intraoperative digital subtraction angiography and intraoperative indocyanine green video angiography during intracranial aneurysm surgery. J. Neurosurg. 1–8 (2018) doi:10.3171/2018.1.JNS172253.

58. Alander, J. T. et al. A Review of Indocyanine Green Fluorescent Imaging in Surgery. Int. J. Biomed. Imaging 2012, 1–26 (2012).

59. Scerrati, A. et al. Indocyanine green video-angiography in neurosurgery: A glance beyond vascular applications. Clin. Neurol. Neurosurg. 124, 106–113 (2014).

60. Sharma, M. et al. The Utility and Limitations of Intraoperative Near-Infrared Indocyanine Green Videoangiography in Aneurysm Surgery. World Neurosurg. 82, e607–e613 (2014).

61. Cavallo, C. et al. Applications of Microscope-Integrated Indocyanine Green Videoangiography in Cerebral Revascularization Procedures. Front. Surg. 6, (2019).

62. de Oliveira, J. G., Beck, J., Seifert, V., Teixeira, M. J. & Raabe, A. ASSESSMENT OF FLOW IN PERFORATING ARTERIES DURING INTRACRANIAL ANEURYSM SURGERY USING INTRAOPERATIVE NEAR-INFRARED INDOCYANINE GREEN VIDEOANGIOGRAPHY. Operative Neurosurgery 61, 63–73 (2007).

63. Chalouhi, N. et al. Safety and Efficacy of Intraoperative Angiography in Craniotomies for Cerebral Aneurysms and Arteriovenous Malformations. Neurosurgery 71, 1162–1169 (2012).

64. Heiles, B. et al. Performance benchmarking of microbubble-localization algorithms for ultrasound localization microscopy. Nat. Biomed. Eng. 6, 605–616 (2022).

65. Rong Li, X. & Jilkov, V. P. Survey of maneuvering targettracking . part I: dynamic models. IEEE Trans. Aerosp. Electron. Syst. 39, 1333–1364 (2003).

66. Xing, P. et al. 3D ultrasound localization microscopy of the nonhuman primate brain. EBioMedicine 111, 105457 (2025).

67. van Heel, M. & Schatz, M. Fourier shell correlation threshold criteria. J. Struct. Biol. 151, 250–262 (2005).

68. Kikinis, R., Pieper, S. D. & Vosburgh, K. G. 3D Slicer: A Platform for Subject-Specific Image Analysis, Visualization, and Clinical Support. in Intraoperative Imaging and Image-Guided Therapy 277–289 (Springer New York, New York, NY, 2014). doi:10.1007/978-1-4614-7657-3_19.

