## Supplemental Information for "4D Ultrasound Localization Microscopy of Deep Cerebral Perforating Arteries for Intraoperative Neurosurgical Guidance"

Supplementary Appendix
Table S1. Patient and Measurement Characteristics.
Table S2. Summary of feasibility.
Figure S1. Lesion location of all patients relative to skull base and circle of Willis.
Figure S2. Montage of annotated perforating arteries for each patient, rendered in the same view as Figure 3.
Figure S3. Vessel annotation pipeline.

Link videos: https://surfdrive.surf.nl/s/6gqtN7pomg54gnR
Video S1. Whole skull rendering of exemplary Patient 4-1.
Video S2. Zoom-in rendering of exemplary Patient 4-1.
Video S3. Whole skull rendering of the pre-resection acquisition of the large tumor case (Patient 2-1), Figure 6.a.
Video S4. Side by side rendering of zoom-in Patient 2-1 (pre-resection) ULM density map overlay with PC-CTA (Figure 6.b) and annotation map (Figure 6.f)
Video S5. Side by side rendering of zoom-in Patient 2-2 (post-resection) ULM density map overlay with PC-CTA (Figure 6.c) and annotation map (Figure 6.g)

Table S1. Patient and Measurement Characteristics

| # | Sex | Age | Tumor location | Tumor Type | Surgical approach | Tumor size (mm) | PC-CTA | Targeted Perforating Arteries | Measurements | Probe placement |
|---|---|---|---|---|---|---|---|---|---|---|
| 1 | F | 60+ | Anterior clinoid process | Meningioma | Pterional | 20 | Pre | LLSA | Post-resection, single-angle | Deep (intradural) |
| 2 | F | 50+ | Sphenoid | Meningioma | Pterional | 51 | Pre | LLSA | Pre-resection, single-angle; Post-resection, single-angle | Superficial (extradural), Superficial (extradural) |
| 3 | M | 35+ | Tectum | Cavernoma | Posterior intrahemispheral transtentorial | 29 | Pre | PCA | Pre-resection, single-angle | Superficial (intradural) |
| 4 | M | 40+ | Cerebello-pontine angle, extended to prepontine cistern, Meckel's cave, and ambiens cistern | Epidermoid cyste | Pretemporal transcavernous | - | Pre | LLSA | Pre-resection, single-angle | Superficial (extradural), deep (extradural - post clinoidectomy) |
| 5 | M | 45+ | Temporal | Cavernoma | Temporal | - | No | AchA | Pre-resection, single-angle | Superficial (extradural) |
| 6 | F | 50+ | Anterior clinoid process | Meningioma | Pterional | 13 | Pre | LLSA | Pre-resection, single-angle | Deep (extradural, post-clinoidectomy) |
| 7 | F | 50+ | Tuberculum sellae | Meningioma | Pterional | 19 | Post | LLSA, ICA-terminus, RAH | Pre-resection, multiple-angle | Superficial (extradural), deep (extradural – post clinoidectomy) |
| 8 | F | 55+ | Cavernous sinus | Meningioma | Pterional | - | Pre | LLSA | Pre-resection, multiple-angle | Superficial (extradural) |
| 9 | F | 60+ | Olfactory groove | Meningioma | Pterional, pretemporal | 25 | Pre | LLSA, MLSA | Pre-resection, multiple-angle | Superficial (extradural) |
| 10 | F | 60+ | Sphenoid inner/outer-ridge | Meningioma | Pterional | 64 | Pre | LLSA | Pre-resection, multiple-angle | Superficial (extradural) |

LLSA: Lateral lenticulostriate arteries, AchA: Anterior choroidal artery, RAH: Recurrent artery of Heubner, MLSA: Medial lenticulostriate arteries, ICA: Internal carotid artery, PCA: Posterior cerebral artery. Age rounded down to the decade. Tumor size dimension is maximum axial diameter. Age displayed in a 5 year range (e.g., 20+ = 20-24 years)

Table S2 Summary of feasibility

| Stage | N Patients | N Acq | Reason for Exclusion |
|---|---|---|---|
| Total enrolled | 10 | 16 | |
| CEUS-ULM feasible | 8 | 13 | Patient 3: Poor acoustic coupling<br>Patient 10: Only see tumor vessel, attenuation (tumor texture) |
| Quantitatively analyzed (anatomy) | 6 | 11 | Patient 1: lack BrainLab tracking<br>Patient 5: lack PC-CTA |
| Quantitatively analyzed (hemodynamics) | 6 | 9 | Patient 4 Acq2: no reliable cardiac cycle segmentation<br>Patient 9 Acq2: no reliable cardiac cycle segmentation |

Acq=Acquisition.

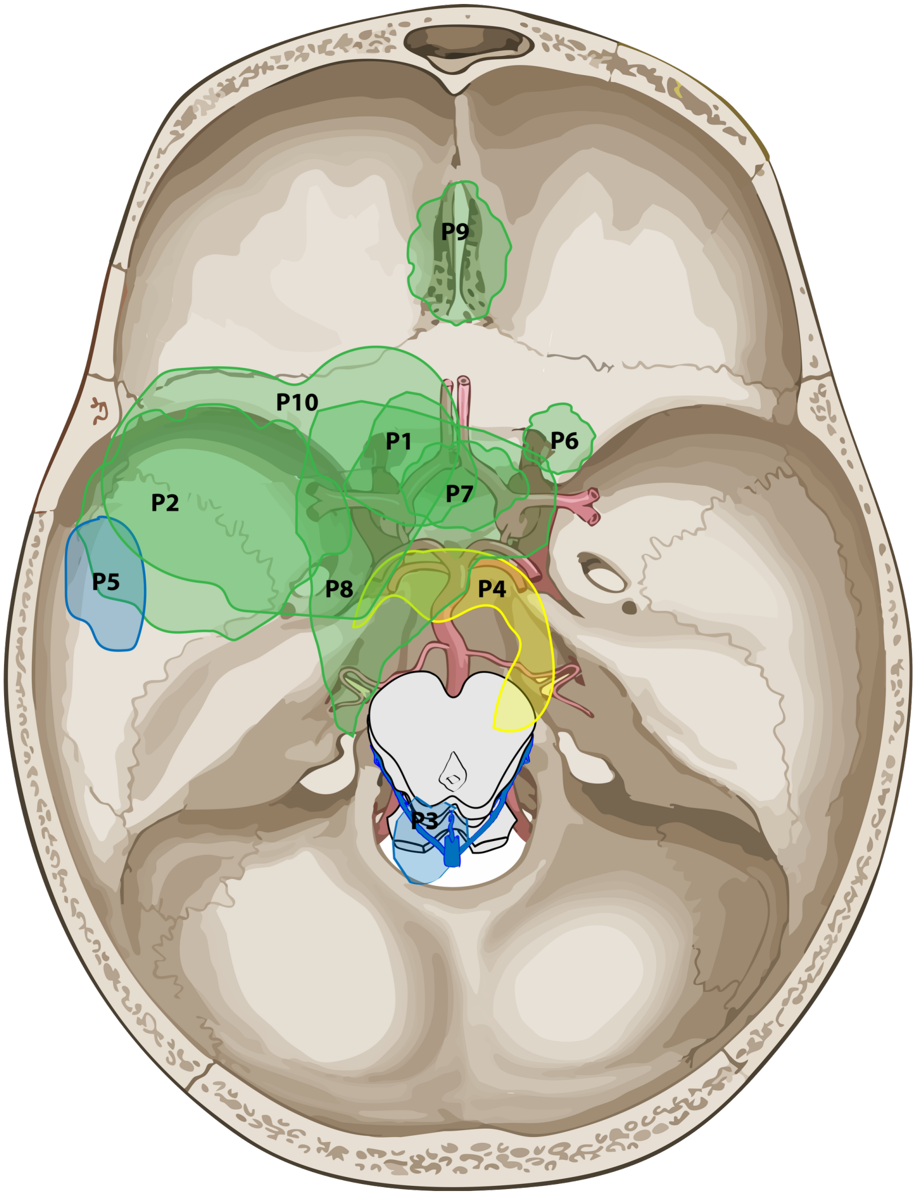


Figure S1. Lesion location of  all patients relative to skull base and Circle of Willis. P-numbers corresponding to patient numbers from Table S1. Green: meningiomas, yellow: epidermoid cyst, blue: cavernomas.

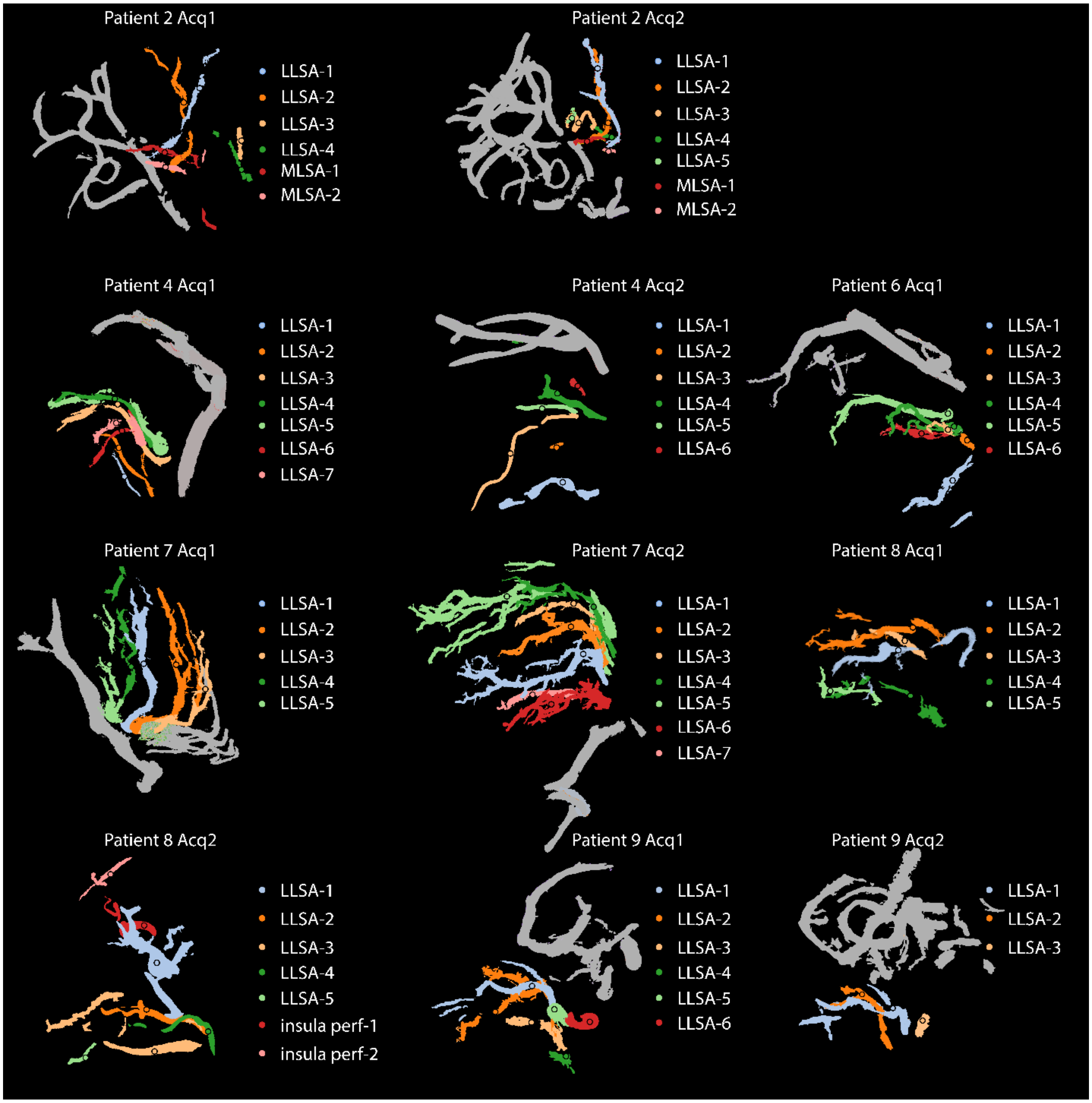

Figure S2. Montage of annotated perforating arteries for each patient, rendered in the same view as Figure 3. LLSA = Lateral lenticulostriate artery; MLSA = Medial lenticulostriate artery. Insula perf = Insula perforator. Numbering of LLSA/MLSA is arbitrary and not consistent between patients or acquisitions.

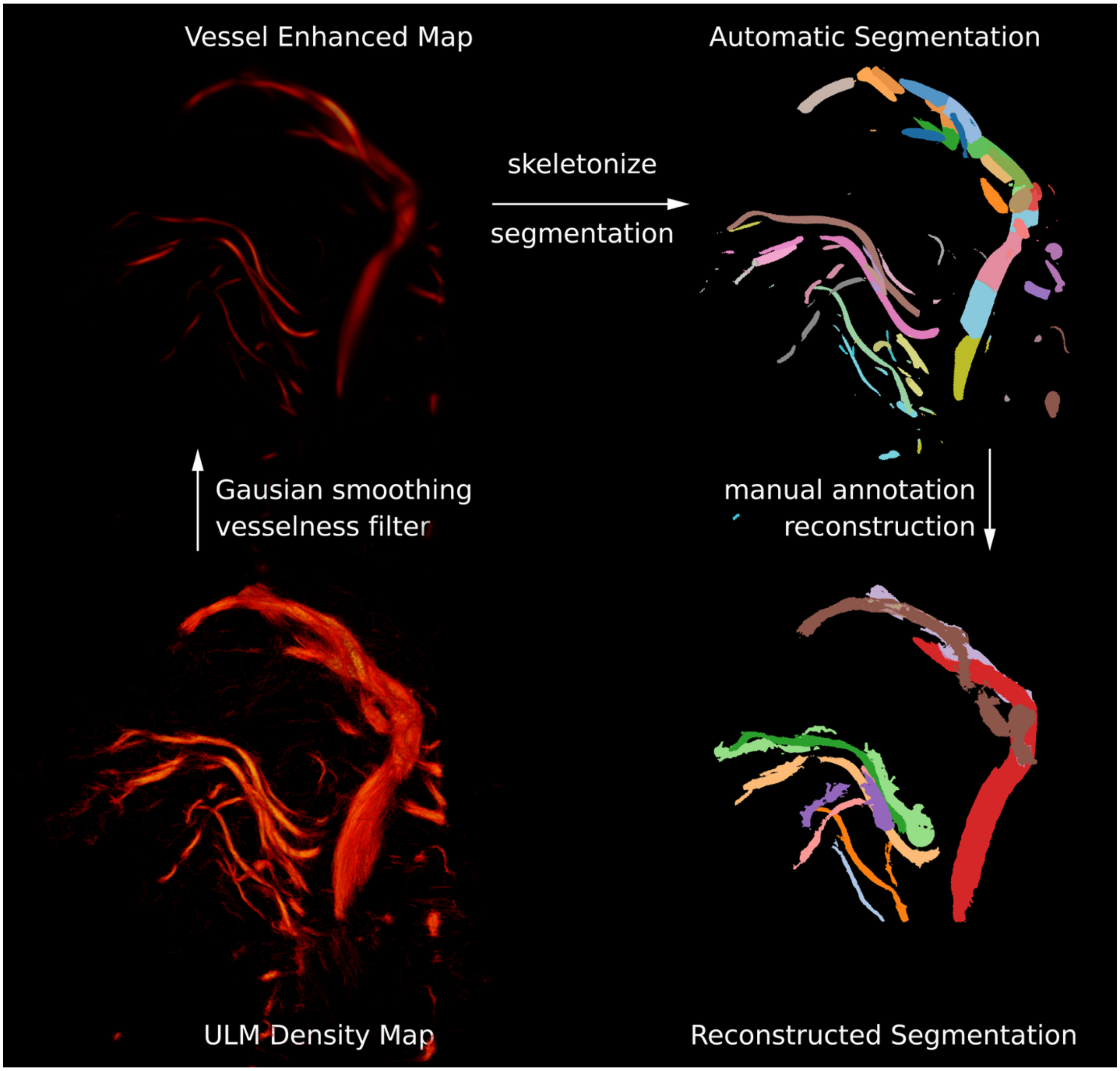

Figure S3. Vessel annotation pipeline. First the ULM density map is smoothed and passed through a Gaussian vesselness filter. Then the smoothed vesselness map is skeletonized and automatically segmented. These automatic segments are then manually reviewed and collated into primary vessels and perforators.
